# Eastern Diet - a healthful dietary pattern from Eastern China: Its characteristics and relation to adiposity, cardiometabolic diseases, mortality, and gut microbiota

**DOI:** 10.1101/2024.07.10.24310112

**Authors:** Yuwei Shi, Juntao Kan, Xinmei Li, Yuji Yu, Changzheng Yuan, Ying Jiang, Qiaoyu Wu, Yufan Hao, Ningling Wang, Wenjie Wang, Emma Yunzhi Huang, Weifang Zheng, Fei Yang, Joyce Wu, April Myers, Ann W. Hsing, Jun Du, Wei He, Shankuan Zhu

## Abstract

**Aim:** The dietary pattern in the downstream Yangtze River region of Eastern China has garnered widespread attention for its potential health benefits but lacks empirical evidence. This study aimed to identify and characterize this dietary pattern, develop a dietary pattern index, and evaluate its associations with adiposity, cardiometabolic diseases (CMDs), mortality, and gut microbiota.

**Methods:** This study used data from 8665 participants aged 18-80 years in the WELL-China cohort (2016-2019, Hangzhou, Eastern China) as the discovery cohort. K-means clustering identified an “Eastern Diet” (EastDiet) pattern and an adherence index based on the 12 food groups and flavor preferences were developed. Adiposity was measured using dual-energy X-ray absorptiometry. Incident CMDs and mortality were tracked through June 2024. Validation involved two external cohorts.

**Results:** The EastDiet pattern was identified characterizing by high plant-based and aquatic food consumption, low refined grains and red meat intake, and a high preference for light-flavored. EastDiet index was favorably associated with lower adiposity. Comparing the highest to the lowest adherence tertiles, hazard ratios (HRs) for CMDs, type 2 diabetes, and cardiovascular diseases were 0.75 (95% CI: 0.62-0.90), 0.76 (0.60-0.96), and 0.71 (0.53-0.94), respectively. All-cause mortality was similarly reduced. Gut microbiota profiles associated with higher EastDiet adherence were linked to improved adiposity and reduced CMDs risk. Validation cohorts replicated these findings.

**Conclusion:** This study identified and characterized the EastDiet pattern from Eastern China and determined its health benefits. Our findings highlighted the potential of the EastDiet as a healthful dietary pattern for Chinese population.

## Introduction

Dietary patterns, representing habitual food combinations, are pivotal to global health and well-being.^1^ These patterns, which include the complex interactions and synergies among various dietary components, provide a holistic view of nutrition beyond the scope of single nutrients or foods.^2, 3^ In China, dietary transitions—from traditional diets rich in whole grains and vegetables to those high in fat and processed foods, have paralleled a surge in cardiometabolic diseases (CMDs).^4–6^ Given China’s significant global population share, identifying a culturally relevant healthy dietary pattern is critical for public health.

Notably, the Yangtze River Delta region, characterized by higher life expectancies and lower rates of obesity and cardiovascular events,^7–9^ offers insight into potentially beneficial regional dietary practices.^10^ This dietary pattern emphasizes lightly flavored and high consumption of vegetables, fruits, aquatic products, soy products, and dairy, which has been recently proposed as a potential healthful model for daily eating across China.^5, 11^ However, this dietary pattern has not yet been identified and lacks empirical evidences for its purported health benefits.

Thus, this study aimed to: (1) identify and define the “Eastern Diet” (EastDiet) in a cohort from Hangzhou, located in the Yangtze River Delta region; (2) develop the EastDiet index; (3) evaluate its associations with adiposity, CMDs including type 2 diabetes (T2D) and cardiovascular diseases (CVD), and all-cause mortality; and (4) investigate gut microbiota signatures reflecting EastDiet adherence and its associations with adiposity and CMDs. These findings were validated in two external cohorts to test their reproducibility and generalizability.

## Methods

### Study population

The present study utilized the WELL-China cohort as the discovery cohort. WELL-China is a population-based prospective cohort that recruited 10268 participants aged 18-80 years from three districts in Hangzhou, Zhejiang Province, China, between 2016 and 2019.^12^ After excluding participants with incomplete or illogical food frequency questionnaire (FFQ) (n = 695), self-reported cancer (n = 205), implausible energy intake (<800 or >4000 kcal/day for males, <500 or >3500 kcal/day for female) (n = 526), missing flavor preference data (n = 14), and missing anthropometry or dual-energy X-ray absorptiometry (DXA) measurements (n = 163), 8665 eligible participants were included in the analysis. For gut microbiota analysis, those without fecal samples or with self-reported severe gastrointestinal disease were further excluded, leaving 6156 participants. A flowchart of participant selection is presented in **Figure S1**. Ethical approval was obtained from the Institutional Review Boards of Zhejiang University and Stanford University, with written informed consent from all participants.

### Dietary assessment and measurement of covariates

Dietary intake was assessed using a validated semi-quantitative FFQ comprising 26 commonly consumed food items in China.^12^ Participants reported the frequency and average amount of each food or beverage consumed over the past 12 months. Detailed descriptions of the FFQ have been published priviously.^13^ Nutritional data were expressed as the energy contribution (%) of each food item.^13, 14^

Baseline covariates were collected through structured face-to-face interviews. Smoking status was categorized as non-smoker, former smoker, and current smoker. Alcohol consumption was categorized as non-drinkers and current drinkers. Physical activity was classified into low, moderate, and high levels using the International Physical Activity Questionnaire-Short Form.^15^ Antibiotic use was defined as described according to previous study.^16^ Fasting venous blood samples were collected after a 12-hour fast to measure fasting blood glucose, triglycerides, and other biomarkers.

### Identification of dietary patterns

Dietary patterns were identified using K-means clustering on standardized dietary intake data (percentage of energy contribution of each food group). The optimal number of clusters was determined using the *NbClust* function. 26 food groups from the FFQ were grouped into 17 food groups based on nutrient similarities and consumption pattern.

### Development of the Eastern Diet index

The “Eastern Diet” (EastDiet) was identified as one of the dietary patterns (Pattern 1 in **Figure 1**). The EastDiet index was developed based on nine healthy food groups (vegetables, fruits, soy products, aquatic products, dairy products, whole grains, tubers/roots, eggs, and nuts), and three unhealthy food groups (refined grains, deep-fried foods, and red/processed meat). Self-reported flavor preference replaced salt intake as a component of the EastDiet, which was not directly collected. Food groups that were inconsistent with health outcomes in previous studies were not included.^14^ Each food group was assigned a score of 1 to 3 based on tertile consumption. Healthy food groups were given positive scores (T1=1, T2=2, T3=3), whereas unhealthy food groups were given reverse scores (T1=3, T2=2, T3=1). Flavor preference was scored from 1 (strong-flavored), 2 (moderate-flavored), and 3 (light-flavored). We summed scores of 12 food groups and flavor preference to derive EastDiet index, ranging from 13 (minimal adherence) to 39 (maximum adherence).

**Figure 1.**
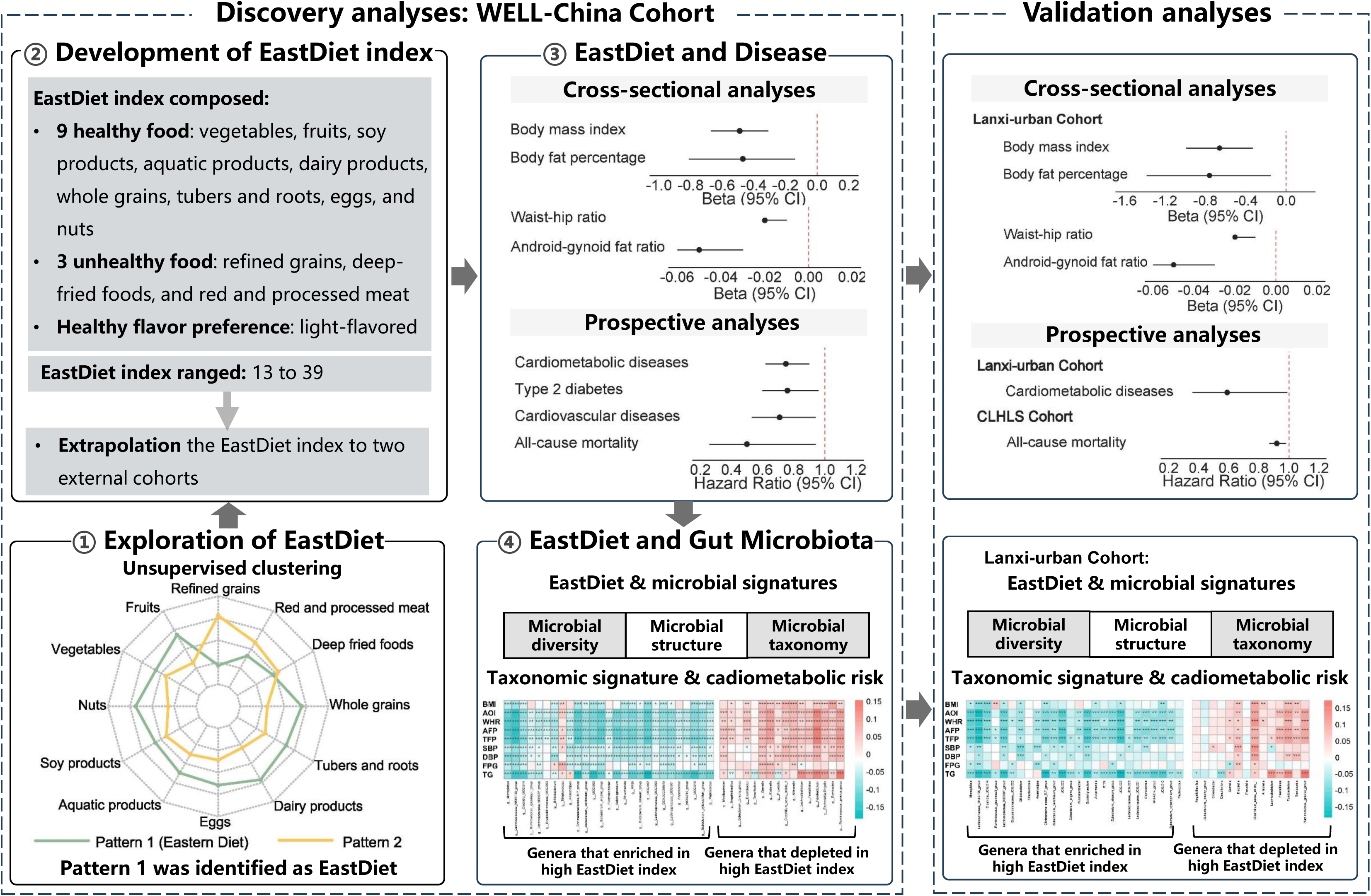
Research overview and key findings. This figure summarizes the study design and main outcomes. Using data from the WELL-China cohort, the Eastern Diet was characterized through unsupervised clustering, leading to the development of the EastDiet index. The EastDiet index was associated with improved adiposity metrics, reduced risks of CMDs, T2D, CVD, and mortality, and healthier gut microbiota profiles. Key findings were validated in two independent cohorts, enhancing the generalizability of the results. The radar plots were generated using standardized data for each food group to enhance visualization. The forest plots were generated using the hazard ratios and beta values comparing the highest tertile to the lowest tertile of the EastDiet index. Abbreviations: EastDiet, Eastern Diet; CMDs, cardiometabolic diseases; T2D, type 2 diabetes; CVD, cardiovascular diseases; HR, hazard ratio; CI, confidence interval.

### Measurement of adiposity-related phenotypes

Detailed information for measuring weight, height, waist, and hip circumference has been previously described^17^. Body mass index (BMI) was calculated as weight (kg) divided by height squared (m^2^). Waist to hip ratio (WHR) was calculated as waist circumference divided by hip circumference. DXA measured fat distribution, including total body fat and body regional fat.^18^ Body fat percentage (BFP) was calculated as total body fat mass divided by weight. Trunk fat mass percentage (TFP), android fat mass percentage (AFP), gynoid fat mass percentage (GFP), and leg fat mass percentage (LFP) were calculated as a percentage of total body fat mass. Android to gynoid fat ratio (AOI) was calculated as android fat mass divided by gynoid fat mass.

### Follow-up for cardiometabolic diseases and mortality

CMDs, including T2D and CVD,^19^ were tracked through electronic linkage to and death registries using International Classification of Diseases, Tenth Revision (ICD-10) codes (T2D: E11/E14, CVD: I20-I23 and I60-I63). Participants were followed from baseline until the onset of CMDs or death or the end of the study period (June 30, 2024), whichever came first.

### Gut microbiota profiling

Fecal samples were collected and stored at −80°C. The V3-V4 region of the 16S rRNA gene was sequenced using the Illumina HiSeq PE-250 platform (Illumina Inc. USA). Detailed information on DNA extraction and paired-end 16rRNA gene sequencing were described previously.^20^ Taxonomic profiles were created using QIIME2.^21^

Pair-end reads were assembled using the qiime tools import command. The DADA2 pipeline was used to filter low-quality regions of the sequences, marker gene Illumina sequences, and chimeric sequences. Reads were then summarized to amplicon sequence variants (ASV) in a feature table and annotated using the Naïve Bayes classifier trained on the Sliva_132 99% OTUs reference databases.

### Validation cohorts

Two external cohorts validated the EastDiet index: Lanxi-urban cohort and Chinese Longitudinal Healthy Longevity Survey (CLHLS). Lanxi-urban cohort was a subset of the Lanxi cohort conducted in urban areas in Lanxi City, Zhejiang Province, China, in 2019.^22^ Using the same exclusion criteria as WELL-China, 2252 participants were included, of which 1788 had available gut microbiota data (**Figure S2**). The EastDiet index (13 components), adiposity, incident CMDs, gut microbiota, and covariates were assessed similarly to WELL-China. CLHLS was a community-based prospective cohort study initiated in 1998 among older adults in China.^23^ This study features a nationally representative sample recruited from 23 provinces across China. We used data from the 2008 wave as a baseline, followed up in the 2011, 2014, and 2018 cycles. After exclusions, 13773 participants were included (**Figure S2**).

Non-quantitative dietary questionnaires provided dietary data, and all-cause mortality was assessed during follow-up. Detailed descriptions of the validation cohorts can be found in the Supplementary Methods section of the Supplementary file.^23–25^

### Statistical analysis

Continuous variables were summarized as means ± standard deviations (SD), while categorical variables were expressed as counts and percentages. For covariates with missing data (less than 1%), a missing indicator approach was applied for categorical variables to minimize data loss.

In the discovery cohort, unsupervised K-means clustering analysis was used to determine dietary patterns. Cross-sectional associations of the EastDiet index with total body fat and regional fat distribution traits were investigated using linear regression, adjusting for age, gender, smoking, alcohol, physical activity, total energy intake, and nutritional supplement use. After excluding the participants with a history of diabetes or cardiovascular diseases at baseline, Hazard ratios (HRs) and 95% confidence intervals (CIs) were calculated to explore prospective associations of the EastDiet index with incident CMDs, T2D, CVD, and all-cause mortality using the Cox proportional regression, adjusting for the aforementioned covariates, and body fat percentage and family history of CMDs, T2D, or CVD.

In gut microbiota analysis, Bray-Curtis dissimilarity was calculated for each participant to determine taxonomy variation. Permutational multivariate analysis of variance (PERMANOVA) was used to quantify the percentage of variance in microbial communities explained by EastDiet and other variables using the Adonis function with 999 permutations. Multivariable-adjusted linear regression models were applied to investigate the associations between the EastDiet index and microbial alpha-diversity (Z-scores).

For per-taxonomic feature tests, taxa with a prevalence below 10% and a relative abundance below 0.01% were filtered out. The relative abundance of the taxa was transformed using rank-based inverse normal transformation. MaAsLin 2 was used to explore genera associated with highest EastDiet level. High-dimensional tests corrected the false discovery rate (FDR) using the Benjamini-Hochberg method with a target FDR of 0.05 considered significant for per-genus test.

To determine whether identified taxonomic features associated with cardiomeatabolic risk, Spearman correlation was used to quantify associations between identified genera and adiposity and other cardiometabolic risk factors, with significance set at 0.05 after the Benjamini-Hochberg correction. Furthermore, an EastDiet-related gut microbiota index (EMI) was developed as a comprehensive representation of the gut taxonomic features. The calculation method was similar to the EastDiet index. Each genus was assigned a score of 1 to 3 based on tertile relative abundance, with higher scores for genera enriched in high EastDiet index and lower scores for genera depleted in high EastDiet index. Multivariable-adjusted linear regression and Cox proportional regression was applied to investigated the prospective associations of EMI with adiposity-related indices and incident CMDs, respectively.

In the validation analysis, the EastDiet index was extrapolated to the Lanxi-urban and CLHLS cohorts. In the Lanxi-urban cohort, multivariable-adjusted linear regression and Cox proportional regression were employed to explore the associations of the EastDiet index with adiposity and CMDs, adjusting for the same covariates as WELL-China. Gut microbiota results of the discovery cohort were replicated. In the CLHLS, Cox proportional regression was used to assess the association between EastDiet index and all-cause mortality, adjusting for the age, gender, ethnic, residence, geographic regions, BMI, smoking, alcohol, physical activity, and history of CMDs. All statistical analyses were performed using Stata 15.0 (StataCorp) and R version 4.2.2. Statistical significance was defined as a two-tailed P-value < 0.05.

## Results

The research overview summary is shown in **Figure 1**. This study identified and characterized the Eastern Diet (EastDiet) pattern and developed an adherence index. The EastDiet was associated with favorable outcomes, including reduced adiposity, lower risks of cardiometabolic diseases (CMDs) and all-cause mortality, and improved gut microbiota profiles. Findings were validated in two external cohorts.

### Identification of Eastern Diet

Among the 8665 participants from the WELL-China cohort, two distinct dietary patterns were identified through K-means clustering (the optimal number of clusters was two), as shown in **Figure 2**. Pattern 1, representing 49.4% of participants, aligned closely with the healthy eating habits described in the 2022 Chinese Dietary Guidelines, including high consumption of vegetables, fruits, soy products, aquatic products, dairy, and a preference for light-flavored foods.^11^ Pattern 1 also captured another seven recognized characteristics of healthy eating behaviors, including high consumption of whole grains, tubers and roots, eggs, and nuts, alongside low consumption of refined grains, red and processed meat, and deep-fried foods.

**Figure 2.**
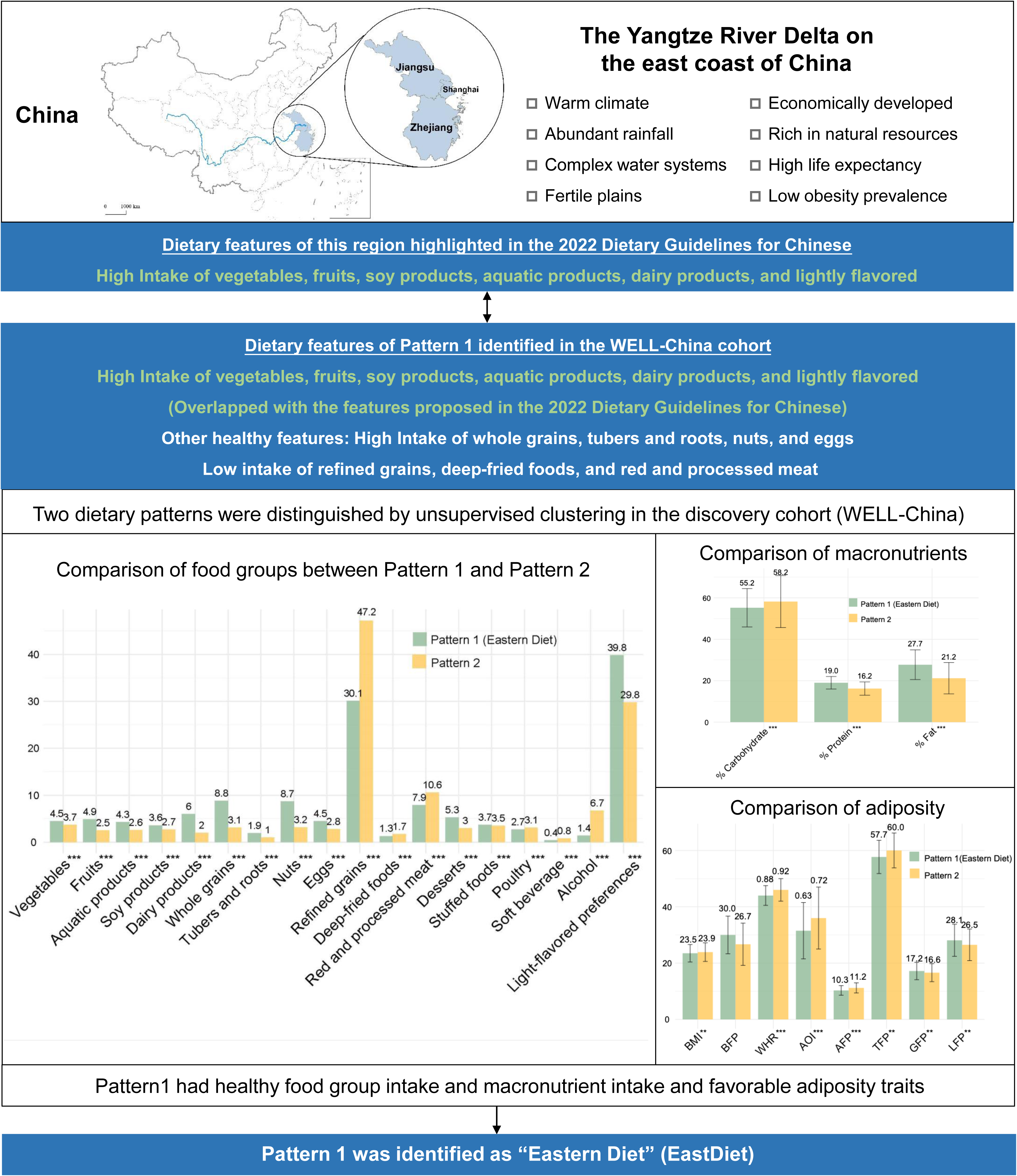
Flowchart of the Eastern Diet identification in the WELL-China cohort. This flowchart outlines the process of exploring and identifying the Eastern Diet. Eastern Diet confirmed the healthy eating characteristics proposed by 2022 Chinese Dietary Guidelines and expanded upon these recommendations. Key differences in nutrient intake and adiposity traits were evaluated using age- and gender-adjusted regression models, highlighting the distinctiveness and health properties of the Eastern Diet. *P*-values were calculated using the age- and gender-adjusted linear regression in the comparisons of the intake of food groups, macronutrients, and adiposity traits, accounting for the significant differences in age (mean: 55.5 vs. 53.2) and gender (79.7% vs. 43.65%) between Pattern 1(EastDiet) and Pattern 2. Significant correlations are indicated with an asterisk.

Compared to Pattern 2, participants adhering to the Pattern 1 had higher protein and fat intake and lower carbohydrate consumption. They also exhibited more favorable adiposity metrics, including lower BMI, AFP, TFP, WHR, and AOI, but higher LFP and GFP, after adjusting for age and gender. Based on these findings, this dietary pattern was designated as the Eastern Diet (EastDiet).

### EastDiet index and participant characteristics

Each participant’s adherence to the EastDiet was evaluated by a 13-dimensional EastDiet index with a possible range from 13 to 39 in the discovery cohort. Participants were stratified into tertiles of EastDiet index scores. Those in the highest tertile were more likely to be older age, female, abstain from smoking and alcohol consumption, lower energy intake and less nutritional supplement use (**Table 1**). These trends were consistent in external cohorts, with participants in the highest tertile of EastDiet adherence sharing similar characteristics (**Tables S1 and S2)**.

**Table 1.** Participant characteristic across tertiles of the Eastern Diet index in the WELL-China cohort.

| Characteristics<sup>a</sup> | Total<br>(N=8665) | Tertile 1<br>(N=3123) | Tertile 2<br>(N=2967) | Tertile 3<br>(N=2575) |
| --- | --- | --- | --- | --- |
| Age (years) | 54.4±13.3 | 51.6±13.8 | 54.6±13.2 | 57.4±12.1 |
| Gender (Female) | 5324(61.4) | 1371(43.9) | 1903(64.1) | 2050(79.6) |
| Smoking status |  |  |  |  |
| Never | 6445(74.4) | 1909(61.1) | 2278(76.8) | 2258(87.7) |
| Former | 611(7.1) | 259(8.3) | 218(7.3) | 134(5.2) |
| Current | 1598(18.4) | 951(30.5) | 466(15.7) | 181(7.0) |
| Current drinkers |  |  |  |  |
| No | 4783(55.2) | 1470(47.1) | 1688 (56.9) | 1625 (63.1) |
| Yes | 3882(44.8) | 1653(52.9) | 1279 (43.1) | 950 (36.9) |
| Total energy intake (kcal) | 1370.4±527.4 | 1419.3±574.0 | 1347.1±514.8 | 1337.8±476.1 |
| Physical activity |  |  |  |  |
| Low | 1411 (16.3) | 655 (21.0) | 445 (15.0) | 311 (12.1) |
| Moderate | 4298 (49.6) | 1391 (44.5) | 1540 (51.9) | 1367 (53.1) |
| High | 2909 (33.6) | 1055 (33.8) | 965 (32.5) | 889 (34.5) |
| Nutritional supplement use |  |  |  |  |
| No | 5236 (60.4) | 2153 (68.9) | 1808 (60.9) | 1275 (49.5) |
| Yes | 3429 (39.6) | 970 (31.1) | 1159 (39.1) | 1300 (50.5) |
| Family history of CMDs |  |  |  |  |
| No | 5995 (69.2) | 2274 (72.8) | 2054 (69.2) | 1667 (64.7) |
| Yes | 2670 (30.8) | 849 (27.2) | 913 (30.8) | 908 (35.3) |
| BMI | 23.7±3.2 | 24.0±3.42 | 23.7±3.2 | 23.3±3.0 |
| BFP | 28.4±7.3 | 26.9±7.60 | 28.7±7.2 | 29.7±6.7 |
| WHR | 0.9±0.1 | 0.9±0.1 | 0.9±0.1 | 0.9±0.1 |
| AOI | 0.7±0.2 | 0.7±0.2 | 0.7±0.2 | 0.6±0.2 |
| TFP | 58.9±6.2 | 59.9±6.3 | 58.8±6.3 | 57.8±5.9 |
| AFP | 10.8±1.8 | 11.2±1.9 | 10.7±1.8 | 10.4±1.7 |
| GFP | 16.9±3.1 | 16.7±3.2 | 16.9±3.2 | 17.2±3.0 |
| LFP | 27.3±5.7 | 26.7±5.7 | 27.4±5.8 | 27.9±5.5 |

Abbreviations: CVD, cardiovascular diseases; BMI, body mass index; BFP, body fat percentage; WHR, waist-hip ratio; AOI, android-gynoid fat ratio; TFP, trunk fat percentage; AFP, android fat percentage; GFP, gynoid fat percentage; LFP, leg fat percentage.
<sup>a</sup> Continuous variables are presented as mean ± standard deviations and categorical variables are presented as N (%).

In the WELL-China, comparing participants born in the Yangtze River Delta with those born in other regions of China demonstrated significantly higher EastDiet index scores (**Figure S3**). In the CLHLS, further comparing participants from the Yangtze River Delta compared with those from other regions of China also showed significantly higher EastDiet index (**Figure S4**). These results provided strong supports for the authenticity of the EastDiet.

### EastDiet adherence and adiposity

In the WELL-China cohort, multivariable linear regression showed that higher EastDiet adherence was significantly associated with lower overall adiposity (BMI and BFP) and central fat distribution (WHR, AOI, TFP, and AFP), as well as higher GFP and LFP compared to lowest tertile (all *P* <0.05) (**Table 2**). These associations were replicated in the Lanxi-urban cohort (all *P* <0.05) (**Table 2**). High EastDiet index was also significantly associated with lower blood pressure, fasting blood glucose and triglycerides levels in the WELL-China (all *P* <0.05) (**Table S3**).

**Table 2.**
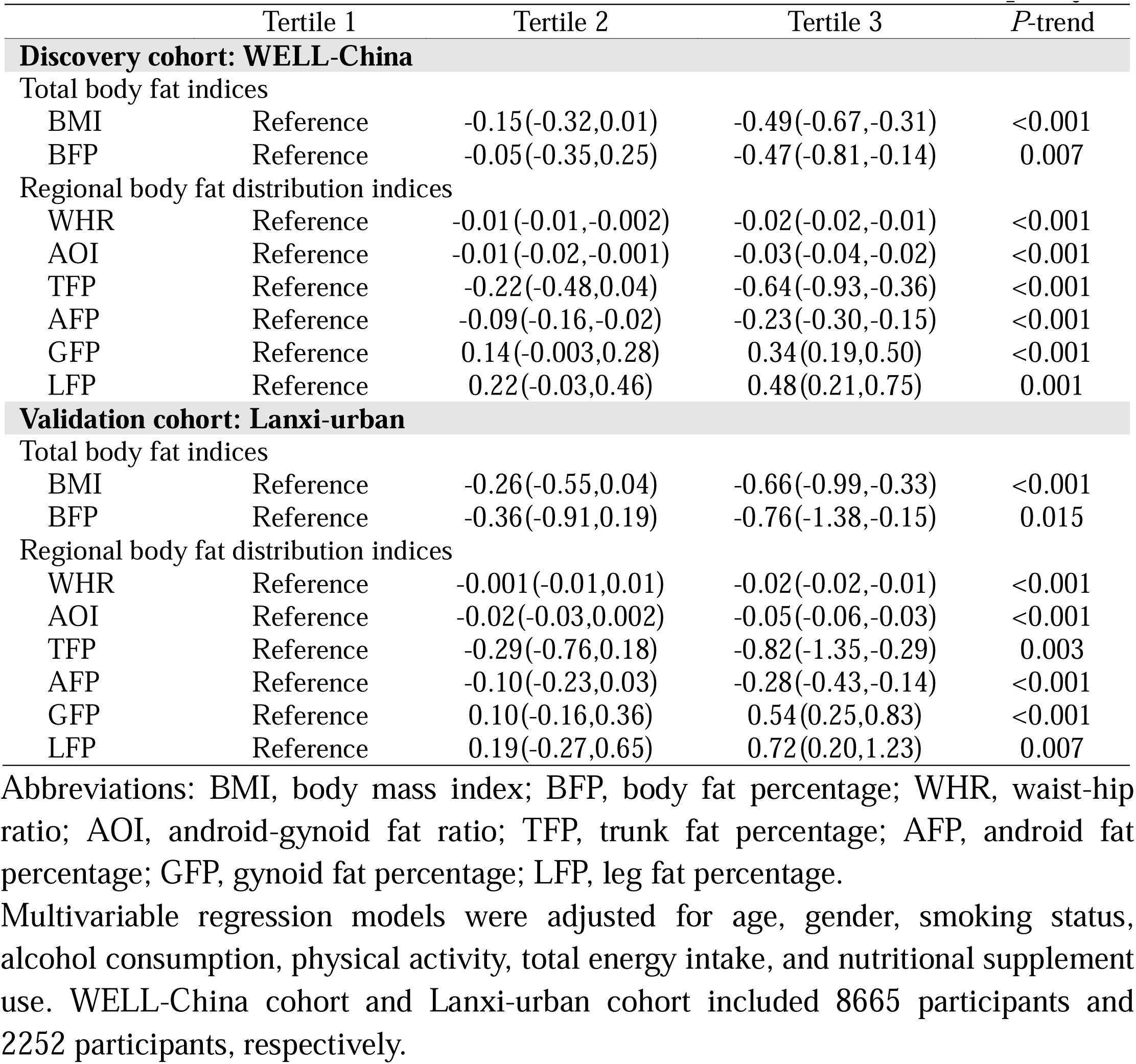
Cross-sectional association of the Eastern Diet index with adiposity.

### EastDiet adherence and cardiometabolic diseases and mortality

In the WELL-China cohort, during a median follow-up of 6.3 years, 762 CMDs were recorded, including 491 T2D and 334 CVD. Additionally, 74 all-cause deaths were documented. After excluding participants with baseline CMDs, EastDiet index was significantly associated with 25% reduced risk of CMDs (tertile 3 vs. 1, HR: 0.75; 95% CI: 0.62-0.90), 24% reduced risk of T2D (HR: 0.76; 95% CI: 0.60-0.96), and 29% reduced risk of CVD (HR: 0.71; 95% CI: 0.53-0.94) (**Table 3**). EastDiet also showed a significant association with all-cause mortality, with an HR (95% CI) of 0.50 (0.26-0.94) for the highest vs lowest EastDiet index categories (**Table 3**).

**Table 3.** Prospective associations of the Eastern Diet index with incident cardiometabolic diseases and all-cause mortality.

|  | Tertile 1 | Tertile 2 | Tertile 3 | *P*-trend |
| --- | --- | --- | --- | --- |
| **Discovery cohort: WELL-China** |  |  |  |  |
| CMDs |  |  |  |  |
| Cases/non-cases | 279/2439 | 263/2254 | 220/2005 |  |
| HR (95% CI) | 1 [Reference] | 0.91(0.76,1.08) | 0.75(0.62,0.90) | 0.003 |
| T2D |  |  |  |  |
| Cases/non-cases | 177/2541 | 171/2346 | 143/2082 |  |
| HR (95% CI) | 1 [Reference] | 0.92(0.74,1.14) | 0.76(0.60,0.96) | 0.024 |
| CVD |  |  |  |  |
| Cases/non-cases | 131/2587 | 108/2409 | 95/2130 |  |
| HR (95% CI) | 1 [Reference] | 0.80(0.61,1.03) | 0.71(0.53,0.94) | 0.017 |
| All-cause mortality |  |  |  |  |
| Cases/non-cases | 35/2683 | 24/2493 | 15/2210 |  |
| HR (95% CI) | 1 [Reference] | 0.70(0.41,1.18) | 0.50(0.26,0.94) | 0.027 |
| **Validation cohort: Lanxi-urban** |  |  |  |  |
| CMDs |  |  |  |  |
| Cases/non-cases | 51/613 | 43/688 | 22/480 |  |
| HR (95% CI) | 1 [Reference] | 0.82(0.54,1.23) | 0.59(0.36,0.99) | 0.043 |
| **Validation cohort: CLHLS** |  |  |  |  |
| All-cause mortality |  |  |  |  |
| Cases/non-cases | 3357/1214 | 3063/1370 | 2970/1462 |  |
| HR (95%CI) | 1 [Reference] | 0.96(0.92,1.01) | 0.92(0.87,0.98) | <0.001 |

Abbreviations: CMDs, cardiometabolic diseases; T2D, type 2 diabetes; CVD, cardiovascular diseases; HR, hazard ratio; CI, confidence interval.
Cox proportional hazard models were adjusted for age, gender, body fat percentage, smoking status, alcohol consumption, physical activity, total energy intake, nutritional supplement use, and family history of CMDs, T2D, or CVD. After excluding participants with a history of diabetes or cardiovascular diseases at baseline, the WELL-China cohort and the Lanxi-urban cohort included 7,460 and 1,897 participants, respectively, with the EastDiet index recalculated accordingly. In the CLHLS, Cox proportional hazard models were adjusted for age, gender, ethnic, residence, geographic regions, BMI, education level, smoking status, alcohol consumption, physical activity, and history of CMDs.

Validation analyses confirmed these associations. The Lanxi-urban cohort showed a 41% reduced risk of CMDs (tertile 3 vs. 1, HR: 0.59; 95% CI: 0.36-0.99) and CLHLS cohort demonstrated a 7% reduced risk of all-cause mortality (HR: 0.93; 95% CI: 0.87-0.98) (**Table 3**).

### EastDiet adherence and gut microbiota

Gut microbiota analyses revealed significant associations with EastDiet adherence. In the WELL China cohort, multivariable analyses of microbial alpha-diversity indicated that high EastDiet index was associated with significantly higher Shannon Index (*P-trend* = 0.028) and Observed features (*P-trend* = 0.030) (**Figure 3A**).

**Figure 3.**
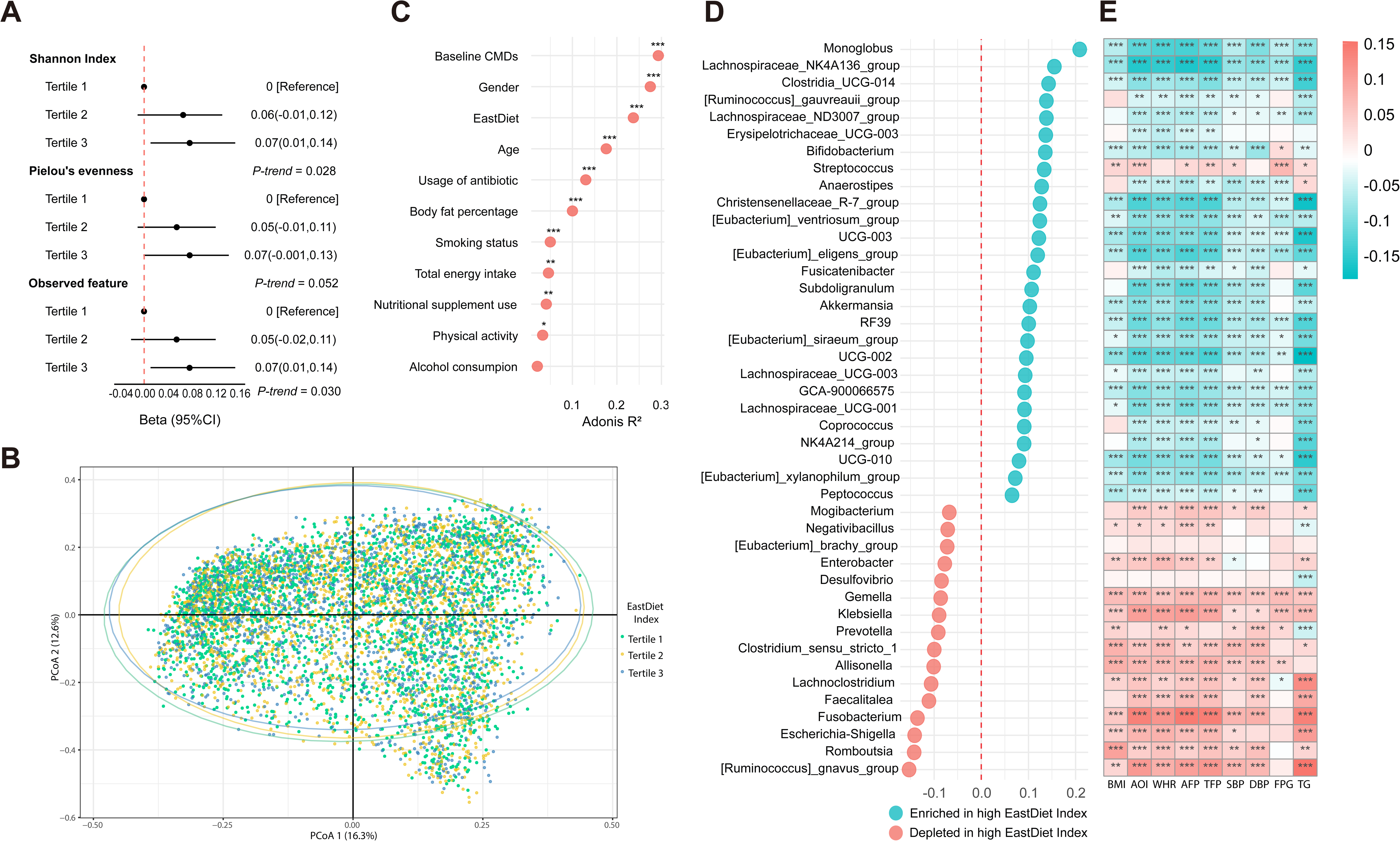
Associations of the Eastern Diet with gut microbial signatures in the WELL-China cohort **A** Multivariable linear regression models were used to evaluate the associations of the EastDiet index with alpha-diversity metrics of the gut microbiota. **B** Principal coordinates analysis (PCoA) based on Bray-Curtis distance was used to assess the dissimilarity across tertiles of the EastDiet index. **C** PERMANOVA test (999 permutations) was used to estimate the proportion of variation in gut taxonomy explained by the EastDiet and other covariates based on Bray-Curtis distance. **D** Multivariate Analysis by Linear (MaAsLin) was used to identify the associations of the EastDiet index with specific microbial genera. **E** Spearman correlation analysis was conducted to assess the associations of the EastDiet-related genera with adiposity and other cardiometabolic risk factors. Covariates included age, gender, body fat percentage, smoking status, alcohol consumption, physical activity, total energy intake, nutritional supplement use, baseline CMDs, and antibiotic use. False discovery rate (FDR) was adjusted using the Benjamini-Hochberg method, with FDR < 0.05 considered statistically significant. Significant correlations are indicated with an asterisk.

Beta-diversity analysis showed significant differences in gut microbial community structure across tertiles of the EastDiet (*P* < 0.001) (**Figure 3B**). The EastDiet index explained 0.24% of the overall gut microbiota structural variance (Adonis R^2^ = 0.24%) (**Figure 3C**). Using MaAsLin 2, 43 genera were significantly associated with the highest tertile of EastDiet (FDR < 0.05) (**Figure 3D**). Among 43 genera significantly associated with EastDiet adherence, 27 were positively correlated. 12 belonged to the *Lachnospiraceae family*, known for fiber fermentation and butyrate production.^26, 27^

Genera that enriched in high EastDiet adherence were inversely correlated with BMI, WHR, AFP, and other cardiometabolic risk factors (**Figure 3E**). Furthermore, a composite EastDiet-related gut microbiota index (EMI) was developed from the identified 43 genera. Higher EMI was significantly associated with the Lower total and central adiposity but higher fat distribution in the legs and hips (all *P* < 0.05) (**Figure 4A**) and 25% reduced risk of CMDs (tertile 3 vs. 1, HR: 0.71; 95% CI: 0.57-0.87) and 38% reduced risk of T2D (HR: 0.70; 95% CI: 0.53-0.91) (**Figure 4B**). These findings suggest a beneficial role of gut microbiota in interacting the health effects of EastDiet adherence.

**Figure 4.**
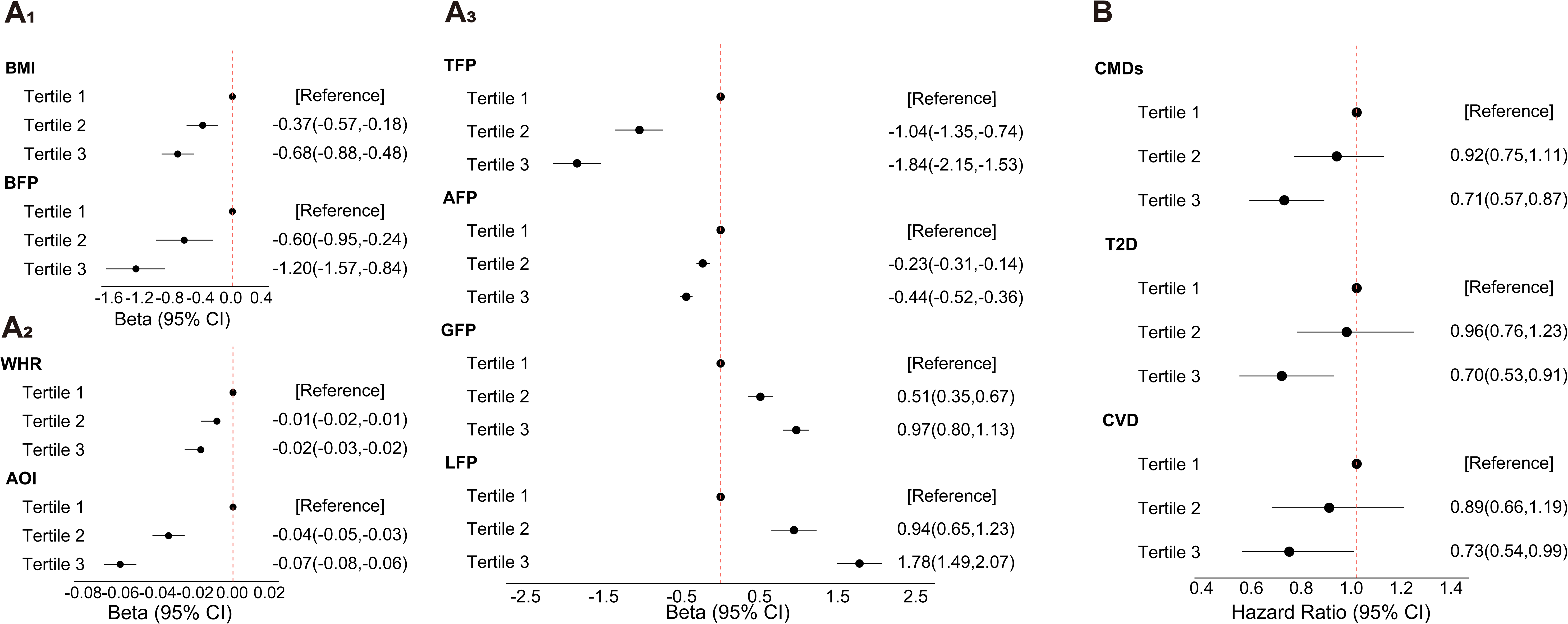
Associations of the Eastern Diet-related gut microbiota index (EMI) with adiposity and incident cardiometabolic diseases in the WELL-China cohort A_1&2&3_ Multivariable linear regression models were used to evaluate the associations of the EMI with adiposity-related indices. B Cox proportional hazard models were employed to assess the associations of the EMI with CMDs, T2D, and CVD. Covariates for the analyses of adiposity indices included age, gender, smoking status, alcohol consumption, physical activity, total energy intake, nutritional supplement use, and antibiotic use. For analyses of incident diseases, body fat percentage and family history of CMDs, T2D, or CVD were additionally included as covariates. Abbreviations: BMI, body mass index; BFP, body fat percentage; WHR, waist-hip ratio; AOI, android-gynoid fat ratio; TFP, trunk fat percentage; AFP, android fat percentage; GFP, gynoid fat percentage; LFP, leg fat percentage; CMDs, cardiometabolic diseases; T2D, type 2 diabetes; CVD, cardiovascular diseases; HR, hazard ratio; CI, confidence interval.

In the Lanxi-urban cohort, beta diversity results replicated those observed in the WELL-China cohort (**Figures S5B and S5C**). Among the 43 identified genera in the WELL-China cohort, 40 genera were annotated and preserved after filtration. 11 genera were significantly and consistently associated with the high EastDiet index (FDR < 0.25) (**Figure S5D**). The similar results on the associations of single genus and EMI with adiposity were observed with those in the WELL-China cohort (**Figures S5E and S6A**). Although a significant association between EMI and CMDs risk was not observed in the Lanxi-urban cohort, trends were consistent with the discovery cohort (**Figure S6B**).

## Discussion

In this large-scale cohort study, we utilized unsupervised clustering to identify and characterize a health-oriented dietary pattern prevalent in the Yangtze River Delta of Eastern China, termed the “Eastern Diet” (EastDiet). This pattern was distinguished by high consumption of plant-based and aquatic food, low intake of refined grains and red meat, and a preference for light-flavored diet. For the first time, we defined the specific components of the EastDiet pattern and developed the EastDiet index to measure its association with health outcomes. Our findings revealed that higher EastDiet adherence was associated with lower total and central adiposity, reduced risks of CMDs, including T2D and CVD, as well as all-cause mortality. Furthermore, gut microbiota signatures reflected adherence to the EastDiet and were linked to improved adiposity and reduced CMDs risks. Validation in two external cohorts confirmed the robustness of these findings.

With one-sixth of the global population, China has a pressing need for culturally tailored dietary patterns that promote health. Unlike the Mediterranean or DASH diets, which are rooted in Western dietary traditions, the EastDiet aligns with regional preferences and culinary practices, making it a more practical and scalable solution for Chinese populations. Our study firstly provides empirical evidence supporting the 2022 Chinese Dietary Guidelines, which propose a prototype healthy diet from the Yangtze River Delta region. By confirming and expanding upon these recommendations, our findings highlight the potential of the EastDiet to serve as a cornerstone of public health strategies aimed at improving dietary quality across China.

While the EastDiet shares similarities with other health-promoting diets, such as the Mediterranean diet and the Healthy Eating Index,^3, 28^ it also incorporates unique features rooted in local dietary practices. For example, the EastDiet emphasizes soy products, aquatic foods including freshwater fishes, and roots and tubers, which are less prominent in Western diets. Additionally, the low consumption of refined grains and deep-fried foods aligns with global recommendations while addressing the risk factors associated with traditional Chinese diets, which are often characterized by a high glycemic index.^29^

Diet is a key factor in obesity development.^30^ Our data showed an inverse association between EastDiet adherence and whole body fat. In line with our findings, a randomized controlled trial of a diet based on Eastern Chinese-style cuisine found that it could achieve weight loss over six months in Chinese adults with pre-diabetes.^31^ In addition to total fatness, we found an inverse association between EastDiet index and lower central fat distribution. Given that Chinese people tend to develop more trunk fat for a given BMI compared to non-Hispanic Whites,^32^ our findings have important implications on strategies to target abdominal fat accumulation for Chinese people.

According to the analysis of the Global Burden of Disease Study, dietary risks was the top attributors to the CVD burden in China^33^. A prior study also found that the shift in diet pattern in China has led to an increase in T2D prevalence and key dietary risk factors fueling this rise included high intake of refined grains, and insufficient intake of whole grains, seafood, vegetables, fruits, nuts, and dairy.^34^ Notably, EastDiet addresses these risk factors and showed the protective associations with new-onset CMDs, T2D, CVD, and all-cause mortality. These associations were consistent with the protective effects of Mediterranean diet and Healthy Eating Index against T2D, CVD, and mortality.^2, 35^ Thus, the EastDiet offers a practical approach to mitigating cardiometabolic diseases and mortality risk in Chinese populations.

Recent studies has underscore the importance of gut microbiota as a marker of dietary quality.^36^ This study revealed that the significant effect of the EastDiet in influencing overall gut microbiome profiles, similar to the effect observed with the Mediterranean diet.^37^ Higher EastDiet adherence was associated with increased microbial alpha diversity, aligning with previous research linking high diet quality to high microbial diversity.^38, 39^ Furthermore, higher EastDiet adherence was associated with the enrichment of beneficial taxa, particularly members of the *Lachnospiraceae family*. These taxa are known for their role in fiber fermentation and butyrate production, which contribute to intestinal homeostasis and cardiometabolic health.^26, 27, 40^ Our results on the relationship between genera and cardiometabolic risk factors also supported this beneficial effect. In addition, the EastDiet-related gut microbiota index (EMI) provides a novel tool for linking dietary patterns to gut microbial composition and CMD risks, highlighting the value of the microbial signature in evaluating diet and disease risks.

Our study has several strengths. Firstly, the EastDiet was identified using unsupervised clustering based on a cohort located in eastern China, ensuring it reflects actual dietary behaviors rather than predefined constructs. Secondly, the EastDiet index and findings were replicated in two external cohorts, including a nationally representative elderly cohort, supporting the generalizability of results. Thirdly, this study integrated cross-sectional, prospective, and gut microbiota analyses, providing a holistic understanding of diet-health relationships.

However, our study has several limitations. Firstly, as to the observational design, residual confounding cannot be entirely ruled out, despite extensive covariate adjustments. Secondly, the semi-quantitative FFQ cannot capture all aspects of dietary intake, particularly salt consumption, However, our study was employed self-reported flavor preference to replace it. Thirdly, the relatively short follow-up period may limit the detection of certain long-term health outcomes, particularly for CMDs subtypes. Finally, CLHLS used non-quantitative dietary questionnaires and lacked several food groups of the EastDiet. This may impact the consistency of findings.

## Conclusion

This study identifies the Eastern Diet, a healthful dietary pattern from Eastern China, which not only aligns with regional culinary preferences and mitigates obesity, T2D, CVD and all-cause mortality, but also has potential to be extrapolated to populations from other regions in China. The Eastern Diet highlights its potential to serve as a local reference for daily healthy eating for Chinese people, similar to the Mediterranean diet for Western populations.

## Data Availability

All data produced in the present study are available upon reasonable request to the authors

## Acknowledgements

Yuwei Shi, Wei He, and Shankuan Zhu designed the study and were responsible for the manuscript. Yuwei Shi analyzed data. Yuwei Shi, Juntao Kan, Jun Du, Wei He, and Shankuan Zhu wrote and revised the manuscript. Yuwei Shi, Xinmei Li, Yuji Yu, Ying Jiang, Qiaoyu Wu, and Yufan Hao created the tables and figures. Shankuan Zhu supervised the whole study. All authors contributed to the critical revisions of the manuscript for intellectual content or collect the data. All authors read and revised the manuscript, and approved the final version.

