## Supplement file for "Eastern Diet - a healthful dietary pattern from Eastern China: Its characteristics and relation to adiposity, cardiometabolic diseases, mortality, and gut microbiota"

**Supplementary file**

**Supplementary Method of Lanxi-urban**

We analyzed data from the Lanxi cohort, a community-based study established in Lanxi City, Zhejiang Province, China. 2698 participants aged 18 to 80 years were recruited from urban areas in 2019. After excluding individuals with incomplete or illogical food frequency questionnaire (FFQ) (n = 186), self-reported cancer (n = 23), implausible energy intakes (<800 or >4000 kcal/day for males, <500 or >3500 kcal/day for females) (n = 131), missing flavor preference data (n = 3), or missing anthropometric or dual-energy X-ray absorptiometry (DXA) measurements (n = 103), a total of 2252 eligible participants were included in the analysis. For gut microbiota analysis, participants without fecal samples or those with self-reported severe gastrointestinal diseases were further excluded, leaving 1,788 participants for evaluation. A flowchart of participant selection is provided in **Figure S2**. The study was approved by the Zhejiang University School of Public Health Medical Ethics Committee, and written informed consent was obtained from all participants.

**Supplementary Method of CLHLS**

**Study Population**

The Chinese Longitudinal Healthy Longevity Survey (CLHLS**)** is a community-based, prospective cohort study launched in 1998 to explore factors contributing to healthy longevity among older adults in China.^21^ The study employs a nationally representative sample drawn from 23 provinces, covering approximately 85% of the Chinese population. Ethical approval for the study was granted by the Biomedical Ethics Committee of Peking University, Beijing, China (IRB00001052–13074).

This analysis used the 2008 cycle as the study baseline, with follow-up assessments conducted in the 2011, 2014, and 2018 cycles. All participants included in the study met the following criteria: who completed follow-up after baseline, provided complete and logical responses to the simplified food frequency questionnaire (FFQ), did not have self-reported cancer, and had no missing data for date of death, height, or weight. After applying these criteria, 13298 participants were included in the validation analysis (**Figure S2**).

**Dietary Assessment and Calculation of the EastDiet Index**

EastDiet adherence was evaluated by calculating the EastDiet index within the CLHLS. Dietary information was collected using a simplified food frequency questionnaire (FFQ), which has been previously validated for reliability and accuracy.^22^ Participants reported the frequency of food consumption. Detailed descriptions of the FFQ have been published elsewhere.^23^ Of the 12 food components originally identified in the WELL-China cohort for the EastDiet, 9 were applicable in the CLHLS: fresh vegetables, fruits, soy products, aquatic products, dairy products, nuts, eggs, red meat, and refined grains.

The EastDiet index was calculated by assigning scores to each food item based on intake frequency. Drawing from established guidelines, each food item was assigned a score ranging from 1 to 5. For refined grains, participants who did not consume refined grains as a primary staple received a score of 5, while those who consumed them were scored inversely (from 1 to 5) based on intake quintiles. For fresh vegetables and fruits, scoring was as follows: ‘almost every day’ (5), ‘quite often’ (4), ‘occasionally’ (2), and ‘rarely or never’ (1). For soy products, aquatic products, dairy products, eggs, and nuts, the scoring was: ‘almost every day’ (5), ‘not every day but at least once per week’ (4), ‘not every week but at least once per month’ (3), ‘not every month but occasionally’ (2), and ‘rarely or never’ (1). Conversely, Red meat was scored inversely to soy products, nuts, aquatic products, dairy products and eggs. Thus, the EastDiet index ranged from 9 (indicating minimal adherence) to 45 (indicating maximum adherence) within the CLHLS.

**Ascertainment of Death**

Mortality data were collected for participants who passed away during the follow-up period (2008-2018). Information on death status was obtained through interviews with close family members. When available, the date of death was recorded from official death certificates; otherwise, it was determined based on reports from close relatives or the local residents’ committee. Survival status and date of death were verified during follow-up surveys, with follow-up duration calculated as the interval between the baseline interview and the date of death. Participants lost to follow-up but still alive were considered censored, with follow-up duration calculated from the baseline interview to the last follow-up.

**Measurement of Other Variables**

Participant characteristics were collected through structured questionnaires in face-to-face interviews under stringent quality control measures. Residence was classified into three categories: city, town, or rural. Ethnic was dichotomized as Han or other ethnic groups. Body mass index (BMI) was calculated by dividing weight (in kilograms) by height (in kilograms) squared. Alcohol consumption was categorized as non-drinkers and current drinkers, while smoking status was classified into three groups: non-smokers, former smokers, and current smokers. Physical activity was assessed as a binary variable (yes or no). A history of cardiometabolic diseases was defined as having at least one self-reported diagnosis of hypertension, diabetes, heart disease, stroke, or other cerebrovascular diseases.

**Table S1 Table 1 Participant characteristics across tertiles of the Eastern Diet index in the Lanxi cohort**

| Characteristics ^a^ | Total  (N=2252) | Tertile 1  (N=799) | Tertile 2  (N=864) | Tertile 3  (N=589) |
| --- | --- | --- | --- | --- |
| Age (years) | 57.0 $\pm$11.9 | 57.0 $\pm$12.4 | 57.0 $\pm$ 11.6 | 56.8 $\pm$11.7 |
| Gender (Female) | 1427 (63.4) | 429 (53.7) | 571 (66.1) | 427 (72.5) |
| Smoking status |  |  |  |  |
| Never | 1800 (79.9) | 577 (72.2) | 705 (81.6) | 518 (87.9) |
| Former | 139 (6.2) | 55 (6.9) | 55 (6.4) | 29 (4.9) |
| Current | 313 (13.9) | 167 (20.9) | 104 (12.0) | 42 (7.1) |
| Current drinkers |  |  |  |  |
| No | 1459 (64.8) | 479 (60.0) | 574 (66.4) | 406 (68.9) |
| Yes | 793 (35.2) | 320 (40.1) | 290 (33.6) | 183 (31.1) |
| Total energy intake (kcal) | 2190 $\pm$591 | 2220 $\pm$ 635 | 2210 $\pm$559 | 2110 $\pm$ 568 |
| Physical activity |  |  |  |  |
| Low | 508 (22.6) | 247 (30.9) | 167 (19.3) | 94 (16.0) |
| Moderate | 1153 (51.2) | 354 (44.3) | 461 (53.4) | 338 (57.4) |
| High | 465 (20.6) | 139 (17.4) | 186 (21.5) | 140 (23.8) |
| Nutritional supplement use |  |  |  |  |
| No | 1996 (88.6) | 741 (92.7) | 745 (86.2) | 510 (86.6) |
| Yes | 256 (11.4) | 58 (7.3) | 119 (13.8) | 79 (13.4) |
| Family history of CMDs |  |  |  |  |
| No | 1697 (89.5) | 619 (93.2) | 636 (87.0) | 442 (88.0) |
| Yes | 200 (10.5) | 45 (6.8) | 95 (13.0) | 60 (12.0) |
| BMI | 23.5 $\pm$3.0 | 23.8 $\pm$3.1 | 23.5 $\pm$3.1 | 23.1 $\pm$2.9 |
| BFP | 30.2 $\pm$7.3 | 29.6 $\pm$7.4 | 30.4 $\pm$7.2 | 30.6 $\pm$7.0 |
| WHR | 0.9 $\pm$0.1 | 0.9 $\pm$0.1 | 0.9 $\pm$0.1 | 0.9 $\pm$0.1 |
| AOI | 0.7 $\pm$0.2 | 0.7 $\pm$0.2 | 0.7 $\pm$0.2 | 0.7 $\pm$0.2 |
| TFP | 59.1 $\pm$5.6 | 60.0 $\pm5.8$ | 59.0 $\pm$5.4 | 58.1 $\pm$5.3 |
| AFP | 10.8 $\pm$1.6 | 11.1 $\pm$1.6 | 10.8 $\pm$1.5 | 10.5 $\pm$1.5 |
| GFP | 16.4 $\pm$2.9 | 16.0 $\pm$3.0 | 16.3 $\pm$2.8 | 16.9 $\pm$2.9 |
| LFP | 26.6 $\pm$5.2 | 25.9 $\pm$5.3 | 26.6 $\pm5$.0 | 27.4 $\pm5$.1 |

Abbreviations: CMDs, cardiometabolic diseases; BMI, body mass index; BFP, body fat percentage; WHR, waist-hip ratio; AOI, android-gynoid fat ratio; TFP, trunk fat percentage; AFP, android fat percentage; GFP, gynoid fat percentage; LFP, leg fat percentage.

^a^ Continuous variables are presented as mean ± standard deviations and categorical variables are presented as N (%).

**Table S2 Table 1 Participant characteristics across tertiles of the Eastern Diet index in the CLHLS cohort**

| Characteristics ^a^ | Total (N=13298) | Tertile1 (N=4433) | Tertile2 (N=4432) | Tertile3 (N=4433) |
| --- | --- | --- | --- | --- |
| Age (years) | 86.6 $\pm$11.5 | 86.5 $\pm$11.3 | 86.6 $\pm$11.5 | 86.8 $\pm$11.8 |
| Gender (Female) | 7543 (56.7) | 2411 (54.4) | 2557 (57.7) | 2575 (58.1) |
| BMI (kg/m^2^) | 20.3 $\pm$3.60 | 19.9 $\pm$ 3.43 | 20.1 $\pm$3.49 | 20.9 $\pm$3.78 |
| Residence |  |  |  |  |
| City | 2124 (16.0) | 292 (6.6) | 523 (11.8) | 1309 (29.5) |
| Town | 2722 (20.5) | 855 (19.3) | 1002 (22.6) | 865 (19.5) |
| Rural | 8452 (63.6) | 3286 (74.1) | 2907 (65.6) | 2259 (51.0) |
| Current smoker |  |  |  |  |
| No | 10861 (81.7) | 3573 (80.6) | 3606 (81.4) | 3682 (83.1) |
| Yes | 2437 (18.3) | 860 (19.4) | 826 (18.6) | 751 (16.9) |
| Former smoker |  |  |  |  |
| No | 8983 (67.6) | 2980 (67.2) | 2998 (67.6) | 3005 (67.8) |
| Yes | 4315 (32.4) | 1453 (32.8) | 1434 (32.4) | 1428 (32.2) |
| Alcohol drinker |  |  |  |  |
| No | 10872 (81.8) | 3688 (83.2) | 3587 (80.9) | 3597 (81.1) |
| Yes | 2426 (18.2) | 745 (16.8) | 845 (19.1) | 836 (18.9) |
| Physical activity |  |  |  |  |
| No | 9622 (72.4) | 3507 (79.1) | 3281 (74.0) | 2834 (63.9) |
| Yes | 3676 (27.6) | 926 (20.9) | 1151 (26.0) | 1599 (36.1) |
| Family history of CMDs |  |  |  |  |
| No | 9626 (72.4) | 3330 (75.1) | 3303 (74.5) | 2993 (67.5) |
| Yes | 3672 (27.6) | 1103 (24.9) | 1129 (25.5) | 1440 (32.5) |

Abbreviations: BMI, body mass index.

^a^ Continuous variables are presented as mean ± standard deviations and categorical variables are presented as N (%).

**Table S3 Cross-sectional associations of the Eastern Diet index with cardiometabolic risk factors**

|  | Tertile 1 | Tertile 2 | Tertile 3 | *P*-trend |
| --- | --- | --- | --- | --- |
| **Discovery cohort: WELL-China** | | | | |
| SBP | Reference | -1.75(-2.52,-0.99) | -2.85(-3.69,-2.01) | <0.001 |
| DBP | Reference | -0.96(-1.48,-0.44) | -1.78(-2.34,-1.21) | <0.001 |
| FPG | Reference | 0.01(-0.05,0.08) | -0.01(-0.08,0.05) | 0.724 |
| FPG without baseline T2D | Reference | -0.05(-0.08,-0.02) | -0.07(-0.10,-0.03) | <0.001 |
| TG | Reference | -0.13(-0.19,-0.07) | -0.15(-0.22,-0.09) | <0.001 |
| **Validation cohort: Lanxi-urban** | | | | |
| SBP | Reference | -1.04(-2.75,0.67) | -1.68 (-3.60,0.24) | 0.080 |
| DBP | Reference | -0.36(-1.29,0.58) | -0.74(-1.79,0.31) | 0.166 |
| FPG | Reference | -0.08(-0.22,0.06) | -0.05(-0.21,0.10) | 0.451 |
| TG | Reference | -0.03(-0.18,0.11) | -0.07(-0.23,0.09) | 0.408 |

Abbreviations: SBP, systolic blood pressure; DBP, diastolic blood pressure; FPG, fasting blood glucose; FPG without baseline T2D, fasting blood glucose without type 2 diabetes diagnosed at baseline; TG, triglyceride.

Multivariable regression models of Beta (95% CI) were stratified by age, gender, body fat percentage, smoking status, alcohol consumption, physical activity, total energy intake, and nutritional supplement use. WELL-China cohort included 8621 participants for SBP, 8622 participants for DBP, 8647 for FPG, for FPG without baseline T2D, and 8650 for TG; Lanxi-urban cohort included 2252 participants for SBP, 2252 participants for DBP, 2246 for FPG, and 2246 for TG;

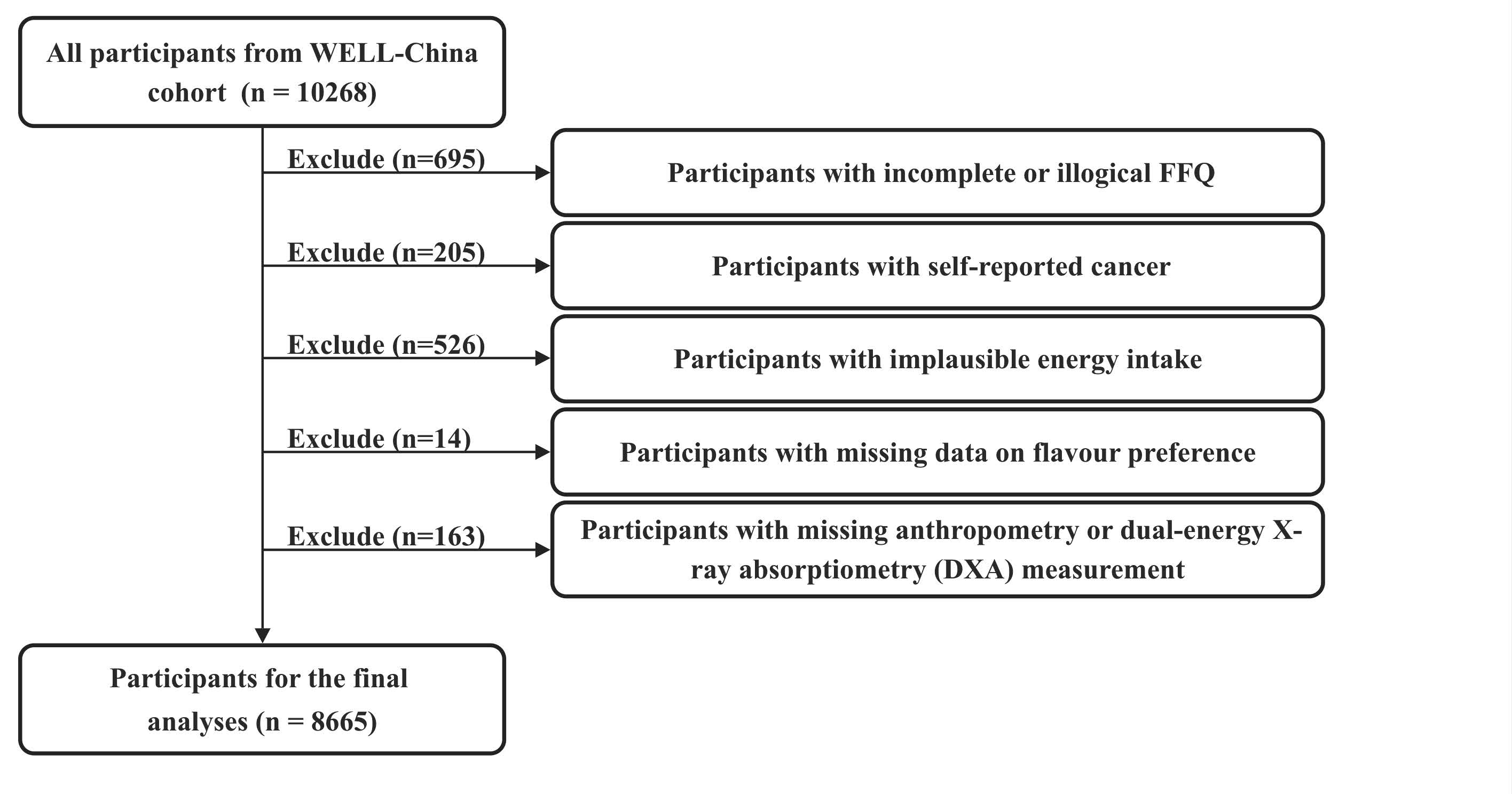

**Figure S1** Flowchart of participant inclusion in the discovery cohort of WELL-China

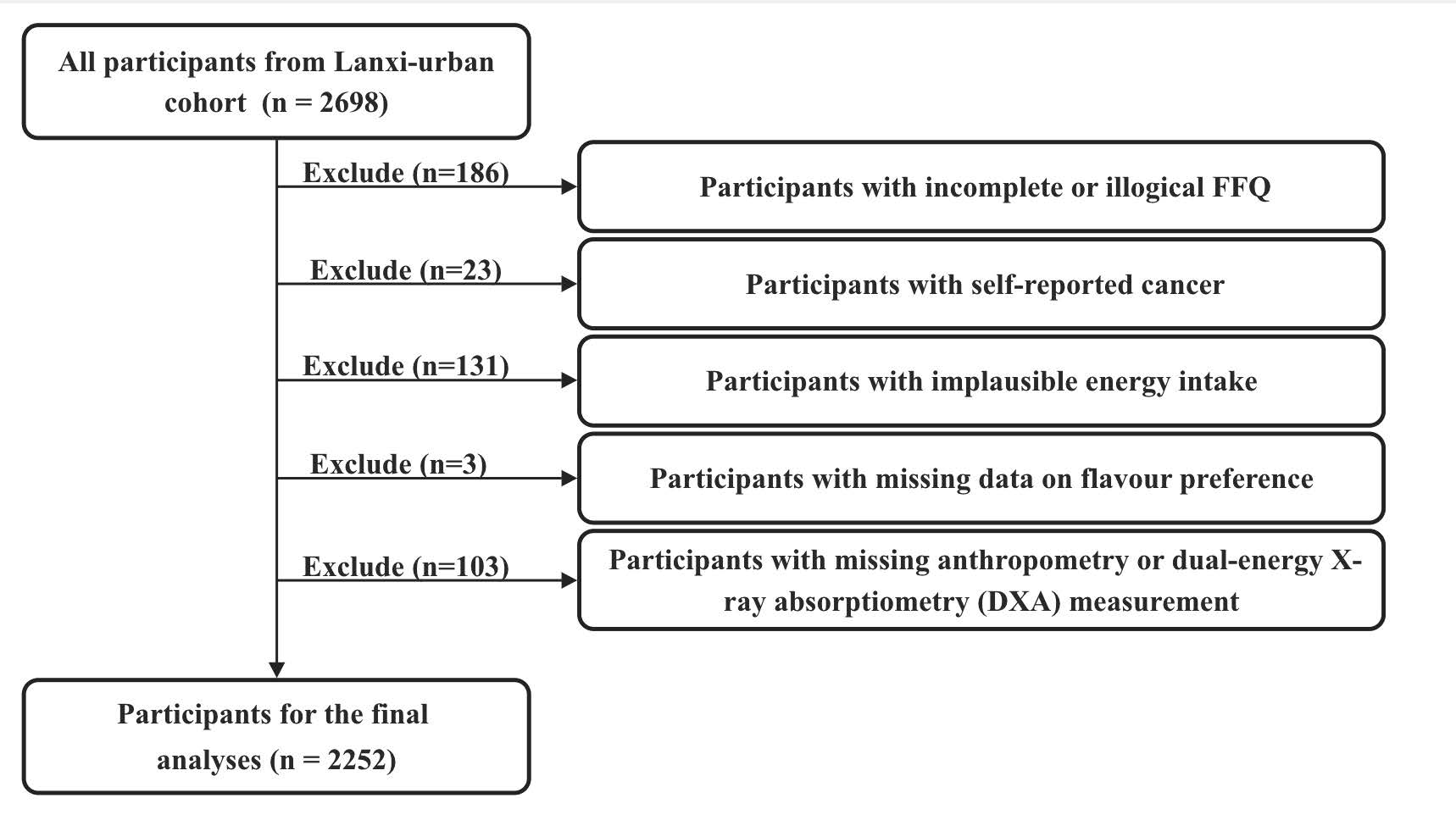

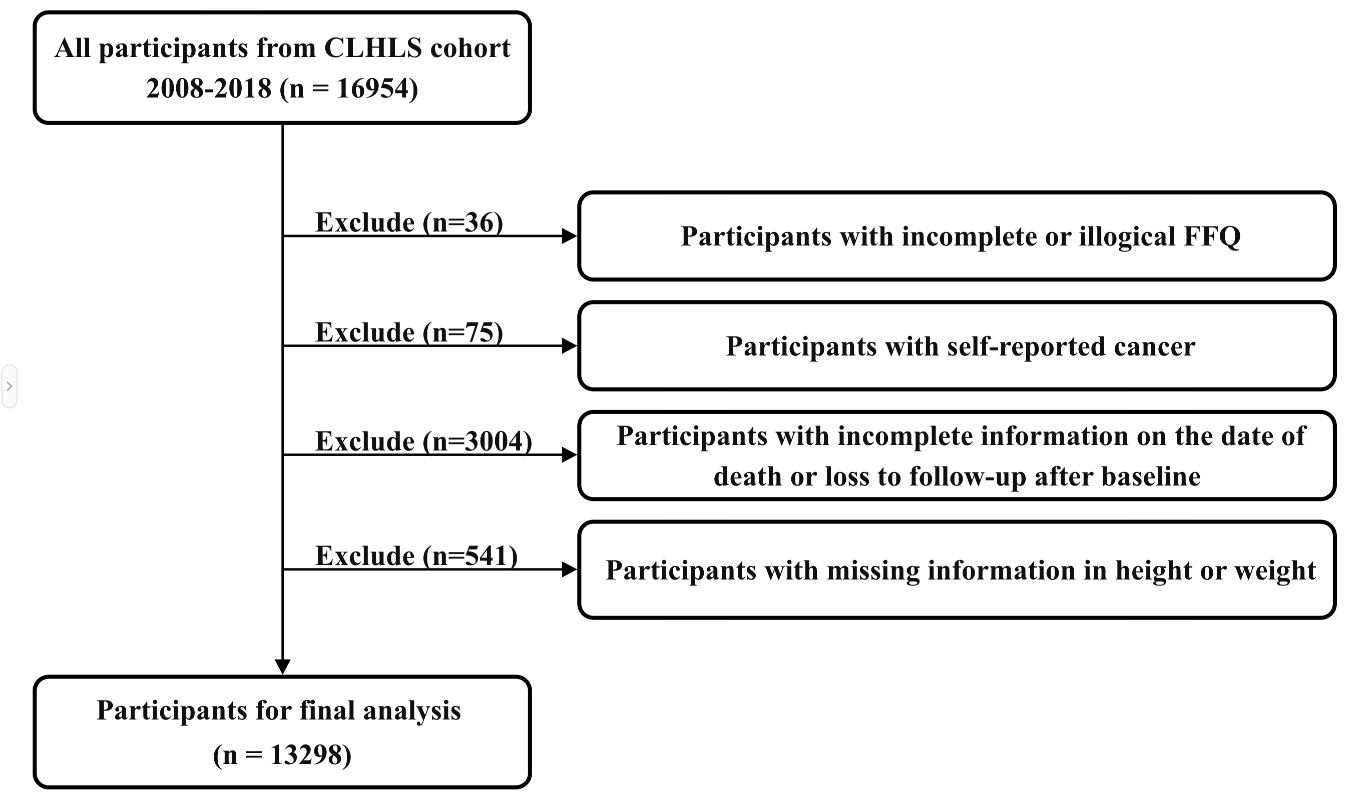

**Figure S2** Flowcharts of participant inclusion in the validation cohorts of Lanxi-urban and CLHLS

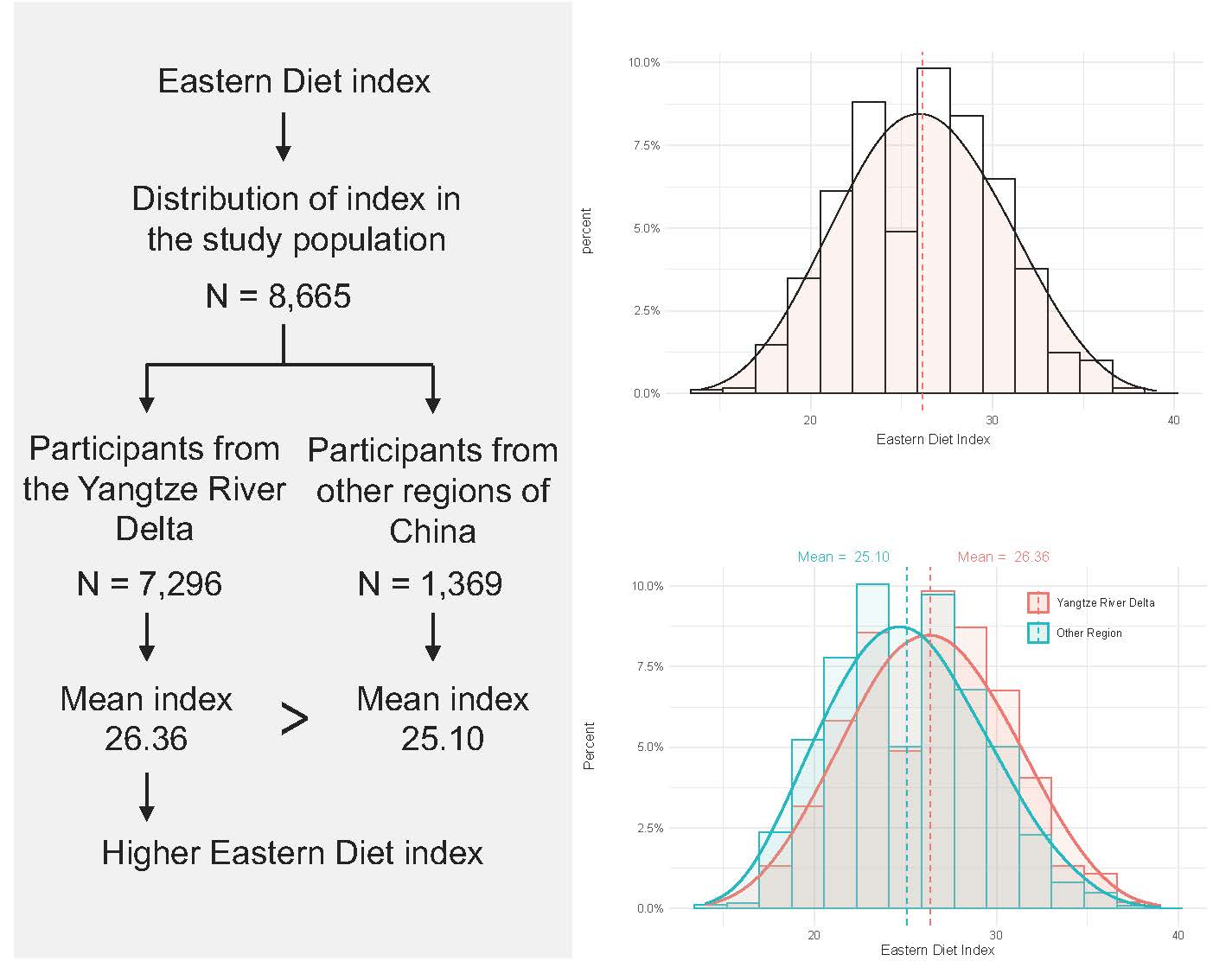

**Figure S3** Distribution and validation of the Eastern Diet index in the WELL-China cohort

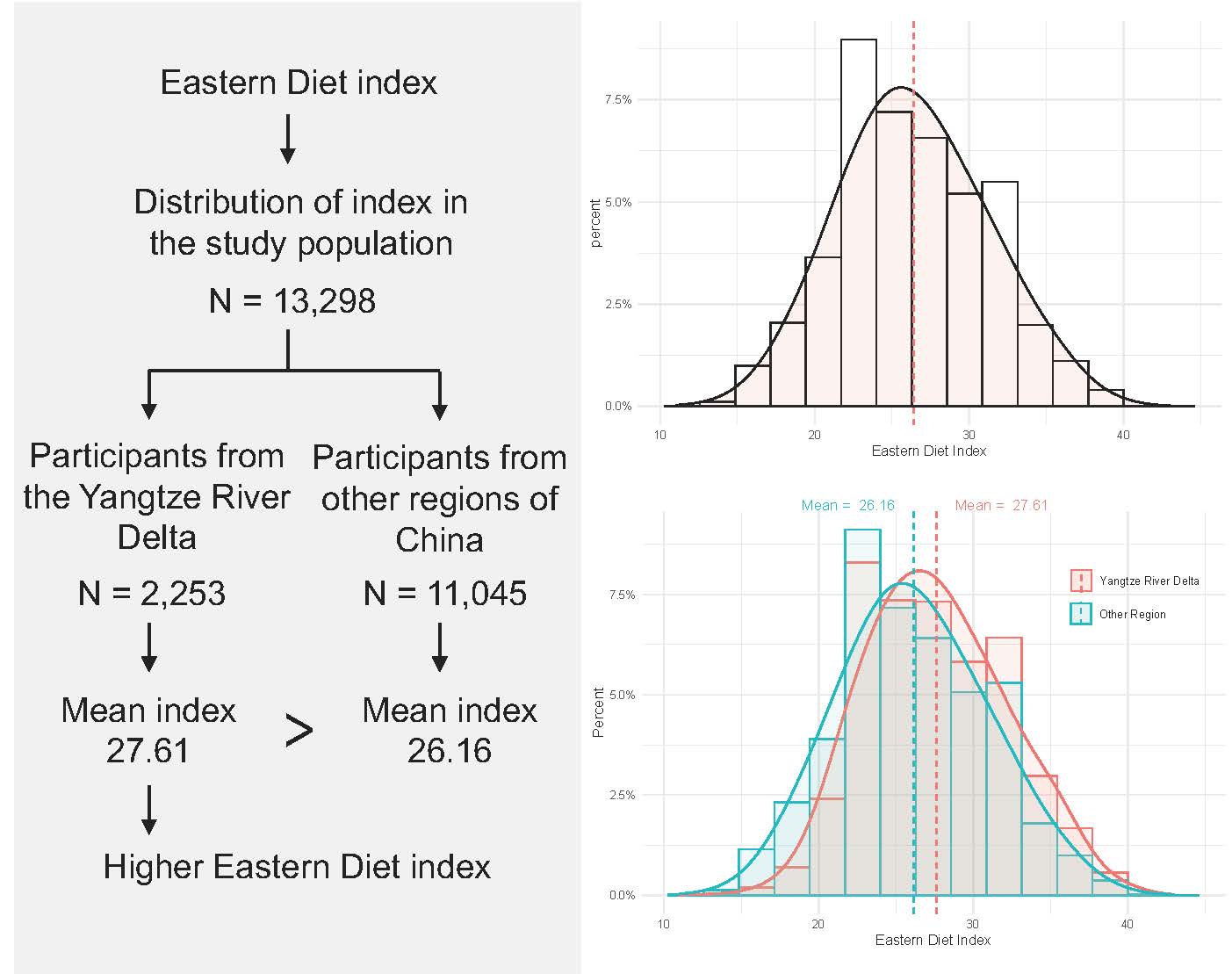

**Figure S4** Distribution and validation of the Eastern Diet index in the CLHLS cohort

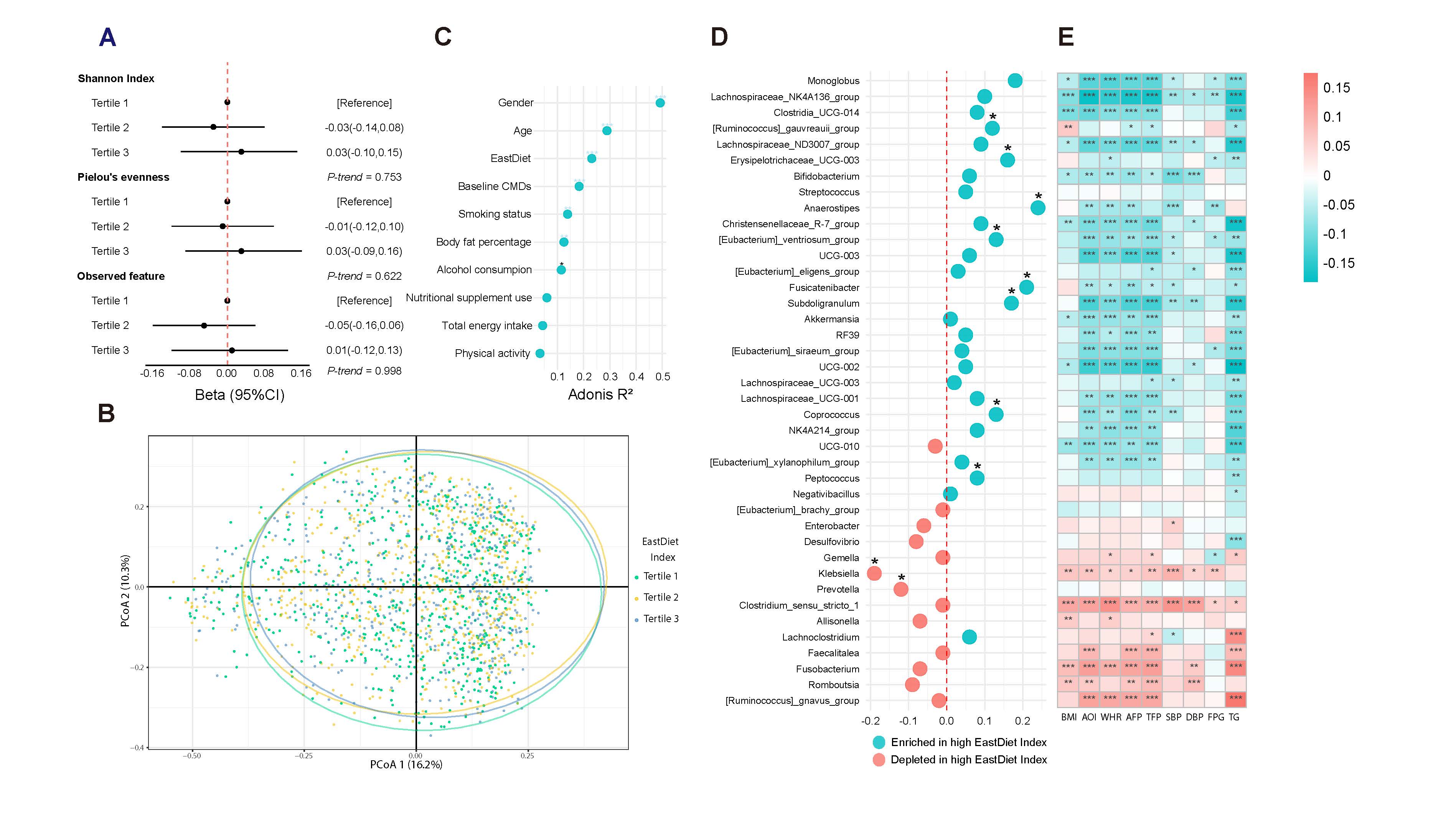

**Figure S5 Validation analysis of the associations of the Eastern Diet index with gut microbial signatures in the Lanxi-urban cohort**

**A** Multivariable linear regression models were used to estimate the associations of the EastDiet index with alpha-diversity metrics. **B** Principal coordinates analysis (PCoA) based on Bray-Curtis distance was used to assess the dissimilarity across tertiles of the EastDiet index. **C** PERMANOVA test (999 permutations) was used to estimate the proportion of variation in gut taxonomy explained by the EastDiet and other covariates based on Bray-Curtis distance. **D** Multivariable-adjusted linear regression was used to replicate the associations of the EastDiet index with identified genera in the discovery cohort. As to this part was a validation analysis with relatively small sample size, false discovery rate (FDR) < 0.25 was considered significant. **E** Spearman correlation was used to estimate the associations of the identified genera with adiposity and other cardiometabolic risk factors. Covariates included age, gender, body fat percentage, smoking status, alcohol consumption, physical activity, total energy intake, nutritional supplement use, and baseline CMDs. FDR was adjusted using the Benjamini-Hochberg method, with FDR < 0.05 considered statistically significant. Significant correlations are indicated with an asterisk.

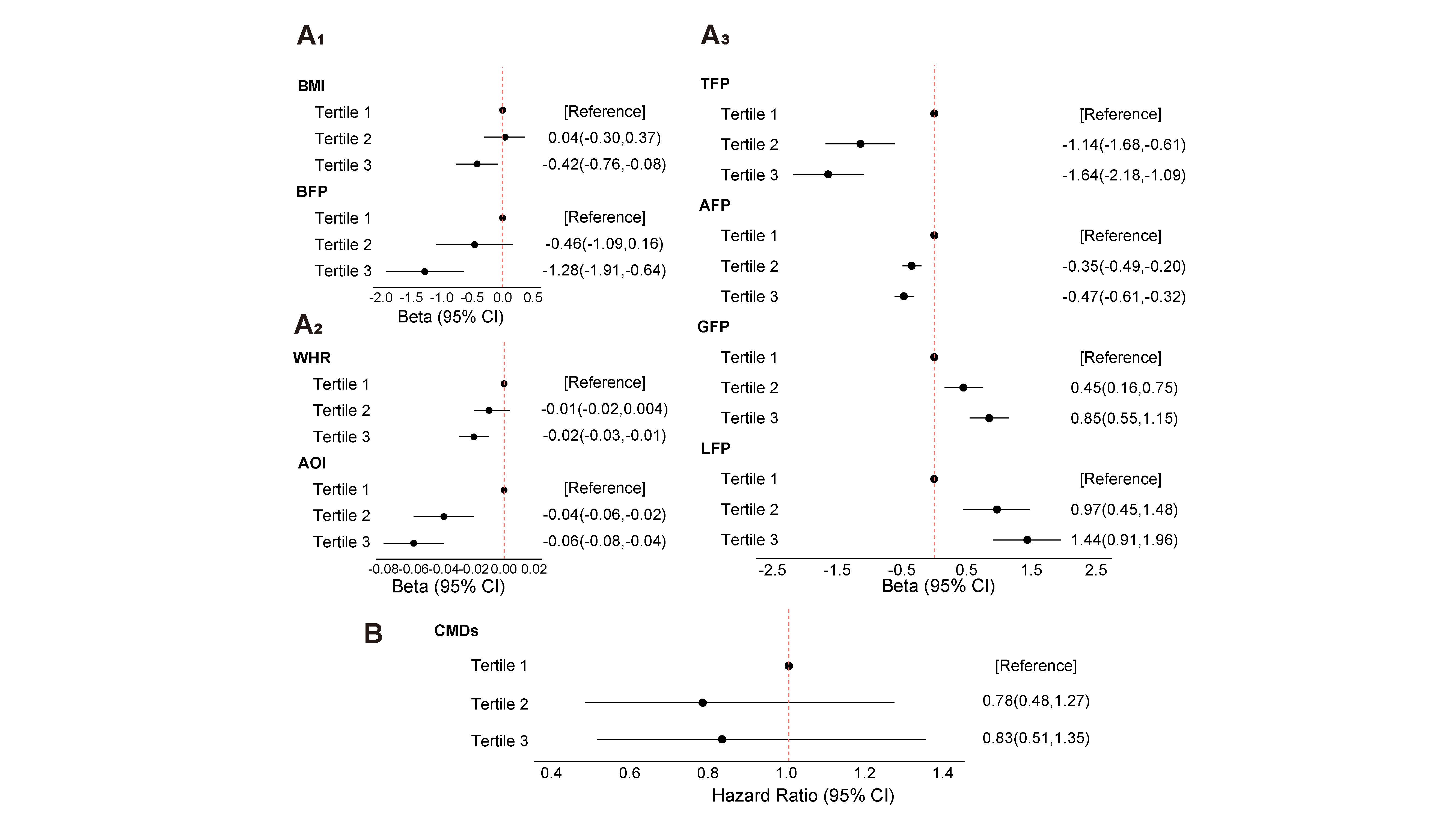

**Figure S6 Validation analysis of the associations of Eastern Diet-related gut microbiota index (EMI) with adiposity and incident cardiometabolic diseases in the Lanxi-urban cohort**

**A_1&2&3_** Multivariable linear regression models were used to evaluate the associations of the EMI with adiposity-related indices. **B** Cox proportional hazard models were employed to assess the associations of the EMI with CMDs. Covariates for the analyses of adiposity indices included age, gender, smoking status, alcohol consumption, physical activity, total energy intake, and nutritional supplement use. For analyses of incident diseases, body fat percentage and family history of CMDs, T2D, or CVD were additionally included as covariates.

Abbreviations: BMI, body mass index; BFP, body fat percentage; WHR, waist-hip ratio; AOI, android-gynoid fat rato; TFP, trunk fat percentage; AFP, android fat percentage; GFP, gynoid fat percentage; LFP, leg fat percentage; CMDs, cardiometabolic diseases; HR, hazard ratio; CI, confidence interval.
